# A ‘Silent Trial’ Assessing the Accuracy of Large Language Models for Assisting Community Health Workers in Low-Resource Settings

**DOI:** 10.64898/2026.02.16.26346409

**Authors:** Natnael Shimelash, Samuel Rutunda, Vaishnavi Menon, Mira Emmanuel-Fabula, Angel Uwimbabazi, Crystal Rugege, Cyprien Nshimiyimana, Ivan Rwema, Mouna Kandekwe, Derbew Fikadu Berhe, Rex Wong, Eric Remera, Emery Hezagira, Jaspret Gill, Lucinda Archer, Richard D. Riley, Alastair K. Denniston, Xiaoxuan Liu, Bilal A. Mateen

## Abstract

Community health workers (CHWs) in low-resource settings deliver variable-quality care. This study used OpenAI’s o3 and Google’s Gemini Flash 2.5 to evaluate whether large language models (LLMs) ‘listening’ to CHW–patient interactions could generate accurate referral decisions. Across 150 participating Rwandan CHWs, 429 encounters were recorded (in Kinyarwanda) and then processed by LLMs. CHWs demonstrated high referral accuracy (97.9% [95% CI: 96.1%-98.9%]), and OpenAI’s o3 performed similarly to CHWs while Gemini 2.5-Flash showed low accuracy (47.3% [95% CI: 42.6%-52.1%]). Assessment of LLM-generated differential diagnoses and management plan quality showed superior performance from o3 compared with Gemini, though both models missed important conditions. In conclusion, the choice of LLM appears to be a critical design decision. Moreover, the high baseline performance of Rwandan CHWs suggests that LLMs are likely to have a limited impact in the current context but could be useful in less well-established CHW programmes. Trial Registration: PACTR202504601308784.

## Introduction

Community health workers (CHWs) are the frontline of Rwanda’s primary healthcare system, delivering preventive, diagnostic, and treatment services within their communities [1,2]. Rapid expansion of Rwandan CHWs’ responsibilities in recent years has significantly increased their workload and introduced unfamiliar clinical scenarios [3–5]. Due to the very limited formal clinical training they receive, their decision-making within those scenarios is guided by structured algorithms that follow rigid, pre-defined logic [6]. These tools are inflexible and do not accommodate complex or atypical presentations well, contributing to delayed recognition and avoidable morbidity [7,8]. Moreover, the need to ensure that the workflow is pragmatic rather than comprehensive, and the turnaround time for revisions to the guidelines – the last formal update having taken place in 2015 [9] – means that as community health needs evolve, there is a risk of over-referring due to issues not being adequately covered by the algorithms [10]. This latter issue is especially important given the Government of Rwanda’s goal to move diagnostic and clinical management services from the clinic to the community.

To strengthen CHW performance, the ‘Community Electronic Medical Record’ (C-EMR), a smartphone-based tool for accessing and editing patient records, as well as making referrals, was recently introduced. While the C-EMR represents progress toward digitization, it largely reproduces existing algorithmic workflows, risking further structurally embedding the aforementioned problems [11,12]. Unlike rule-based tools, large language models (LLMs) can more flexibly interpret nuanced symptoms and adapt to diverse presentations, potentially enhancing CHWs’ decision-making and reducing unnecessary referrals. Across a variety of settings, these tools have shown promise in diagnostic reasoning [13,14], triage [15,16], and treatment planning by synthesizing unstructured information, and generating context-specific recommendations [17]. The prevailing paradigm for LLM-based clinical decision support systems is based on the premise that the health worker prompts an LLM directly (or indirectly). Alternatively, ambient listening solutions – where the LLM ‘listens’ to input without being prompted directly – have demonstrated significant value in supporting administrative tasks such as documentation [18–20], but there is no equivalent evidence for their use as a data-capture solution in combination with an LLM-based clinical decision support system (CDSS).

Whilst some in-silico benchmarking data does exist to evaluate the performance of LLMs in African primary care contexts (and specifically Rwanda) and to do so in local African languages (e.g., Kinyarwanda) [21,22], actual field assessments of their safety, accuracy, and contextual appropriateness in community-level care are missing [23,24]. Thus, leveraging the ongoing digitization agenda in Rwanda, we sought to evaluate whether an LLM-based ‘ambient listening’ solution, which listened to and processed conversations in Kinyarwanda, could assist CHWs to make better clinical decisions.

## Methods

### Design

A prospective observational diagnostic test–evaluation study (i.e., a ‘silent trial’ [M1]) was used to assess the accuracy of referral decisions made by CHWs, OpenAI’s o3, and Google’s Gemini 2.5 Flash. The full protocol is available elsewhere [M2].

### Setting

The Nyabihu and Musanze districts, North-West Rwanda, were selected for their geographic and sociodemographic diversity and for differing stages of CHWApp digital tool adoption: Nyabihu has over 900 CHWs using CHWApp, while 150 CHWs in Musanze recently started using the CHWApp. Together, these two districts encompass both rural and semi-urban populations, capturing contexts relevant to national CHW workflows.

### Data Collection Tool

The Mbaza application (Extended Data Figure 1) was developed for this study by Digital Umuganda and operated silently in the background to audio record the full consultation following consent.

Prior to the launch of the trial, the app was designed based on user experience testing and feedback interviews with CHWs. These sessions explored linguistic comprehension, usability, and interaction preferences for the application. The results emphasized the importance of having the application in Kinyarwanda. Several additional insights related to the LLM outputs, such as a preference for voice over text and the need to improve Kinyarwanda fluency while reducing English code-switching, were integrated over several rounds of prompt engineering and instruction design for the LLMs, which led to the prompt described later.

The Mbaza application was used to capture audio (using the phone’s in-built microphone), generate transcripts, collect structured demographics & clinical information, store pictures of counter referrals, and included a brief user experience survey.

### Participant Recruitment: Inclusion/Exclusion Criteria & Consent

Two sets of participants were recruited in sequence: first, the CHWs, and second, the patients whom they attended to during the study period.

#### Community Health Workers (CHWs)

After ethical approval was obtained, RBC provided a list of all health centers in the two districts to facilitate identification of eligible CHWs (i.e., those who had been provided with smartphones by RBC and routinely used the CHWApp). Random sampling of CHWs was conducted using the RAND function in Excel. Selected CHWs were contacted by research assistants via telephone, during which the study was explained, inclusion and exclusion criteria were checked, and the consent information was read.

Inclusion criteria were completion of the RBC CHW curriculum, ≥3 months of active service at the time of recruitment, and the ability to communicate in Kinyarwanda or English. CHWs were excluded if they had been out of service for >1 month or planned to travel/relocate during the study period.

CHWs who agreed to participate were scheduled for an appointment at their local health centre to complete written informed consent. After consent, the study team installed the Mbaza research app on CHW phones and provided training on the app’s use, patient inclusion criteria, study procedures, and ethical considerations.

#### Patients

All patients presenting to participating CHWs with a new health-related complaint during the study period were considered for the study. After receiving a detailed explanation of the study’s purpose and procedures, patients were invited to participate. For those who agreed, written consent for recording the consultation was obtained before enrollment.

Patients were eligible if they were physically present (i.e., it was not a tele-consultation), and able to provide informed consent. Exclusions included routine visits (for example, routine vaccination, nutritional follow-ups, routine antenatal care), cases requiring immediate emergency care (for example, major trauma), those unable to communicate verbally, consultations where the environment precluded audio recording, or patients who did not speak Kinyarwanda or English.

#### Data Collection, Management & Processing

Once the patient (or the caregiver for those under 18 years of age) provided signed informed consent, the CHW activated the Mbaza app to record the conversation throughout the consultation. After each consultation, the CHW administered a user experience survey to understand the patient’s experience of having their consultation recorded.

Follow-up visits occurred on day 3 as part of routine care. During this visit, CHWs collected information on any changes in health status and subsequent healthcare interventions using a questionnaire embedded in the Mbaza app. Specifically, CHWs collected: patients’ recovery status (recovered/still symptomatic), and details of further action taken regarding the presenting complaint after the initial encounter. If patients required continued monitoring/follow-up, the CHW would record the same information in the Mbaza app on day 14, when they returned. Patients who reported completed symptom resolution by day 3 were not followed up on day 14. If patients were referred to health centres, the CHW would use Mbaza to take a photo and upload the counter referrals detailing the diagnosis and management plan during the follow-ups.

#### Model Selection

Models were selected based on performance in previous study benchmarking LLM performance in a Rwandan healthcare setting [21] which identified gemini-2.0-flash-exp as the leading non-reasoning model and o3-mini-high as the leading reasoning model. Based on these results, we used the most recent versions of the respective models available at the start of the study: Google’s gemini-2.5-flash as the non-reasoning model and o3 (2025-04-16) as the reasoning model. During the course of the study, newer versions were released, and thus, for the sake of generating the most up-to-date results, we sought to replicate the primary analysis of referral decision accuracy (described in detail later) using these more recent releases: gemini-3-pro, and GPT-5.1.

#### Data Processing

Recorded audio files were first reviewed by the research team to assess eligibility, completeness, and audibility. Recordings that met quality criteria were transcribed to Kinyarwanda using a locally developed speech-to-text model, then annotated and anonymized to remove personal identifiers. All transcripts were validated by a team of five annotators and four translators to ensure translation fidelity and de-identification standards.

All cleaned and de-identified transcripts were submitted to two LLMs: OpenAI o3 and Gemini-2.5-Flash, with structured prompts to generate outputs (see ‘Supplementary Material: Prompts’). For each consultation, the LLMs produced three structured outputs: (1) referral decision (refer/do not refer), (2) a ranked list of five differential diagnoses, and (3) a suggested management plan. All outputs were generated after the fact, encrypted, and stored securely. In other words, the LLM outputs were never visible to CHWs during the study.

#### Outcomes and Assessment Procedures

An independent evaluation panel of 12 general practitioners, each registered with the Rwanda Medical and Dental Council and with at least two years of clinical experience, were recruited and trained on the objectives of the study and evaluation tool. The expert panel was used to generate the ‘gold standard’ reference result against which the CHWs and LLMs were compared, suggest a reference diagnosis, and rate the differential diagnoses and management outputs of the LLMs. Panel members received standardized training on the study protocol and evaluation rubric. Four sub-panels of three general practitioners were formed. Cases were randomly assigned to the panels in sequential rotation.

Within each panel, three evaluators first independently assessed the assigned transcripts. Based on the independent assessments, if there was not full agreement in any of the panel responses, the group convened to discuss the case. If consensus among the three evaluators could not be achieved after discussion, a fourth evaluator was appointed to arbitrate and made the final decision based on the arguments put forward by the three original evaluators.

#### Primary Outcome and Estimands

The primary outcome was referral appropriateness. This was measured by comparing each decision to the panel’s “gold standard” reference, which was based solely on the day-0 consultation transcript (reflecting what was known at the time of the initial encounter). The estimands (statistical measures) of interest for the referral appropriateness were the accuracy (% of correct classifications), recall (sensitivity), precision (positive predictive value), specificity, and negative predictive value (NPV).

#### Hindsight referral appropriateness

Hindsight referral appropriateness was assessed using the panel’s revised ground-truth decisions, which considered follow-up information from day-3 and/or day-14 visits as an expanded reference standard. This outcome was reported as hindsight panel judgment: Correct referral decision, Missed referral, and unnecessary referral.

#### Diagnostic Reasoning & Management Plans Assessment

For each case the panel reviewed, they were asked to determine a single most-likely diagnosis. The differential diagnoses lists generated by o3 and Gemini 2.5 Flash were compared with the reference diagnosis made by the panel to calculate the diagnostic accuracy. Concordance is reported as: 1) Top-1, where the reference diagnosis matched the first-ranked diagnosis on the LLM lists; 2) Top-5, where the reference diagnosis appeared anywhere in the LLMs list; and 3) Not listed, where the reference diagnosis did not appear in the list.

The panel also assessed the quality of the differential diagnoses and management plans that were generated by the two LLMs. The quality of diagnoses was assessed in 5 areas:

1. Alignment to clinical standards;
2. Inclusion of relevant conditions associated with the reported symptoms (referred to as recall)
3. Logical reasoning behind the selection of differentials (referred to as reasoning)
4. Exclusion of any critical conditions (referred to as critical omissions); and,
5. Inclusion of irrelevant conditions that potentially detract from diagnostic accuracy (referred to as relevance)

Each of the five areas was assessed using a 5-point Likert scale, with one being the worst score, and five being the best.

Finally, the quality of the LLM proposed management plans were assessed in five areas:

1. Alignment to clinical standards;
2. Omission of crucial elements that are essential to safe and effective care;
3. Bias that could affect applicability;
4. Possible harm;
5. Contextual appropriateness of instructions.

Again, each of the five areas was assessed on a 5-point Likert scale, with 1 being the worst and 5 the best.

#### Patient Experience Queries

Patients’ experience of being recorded was assessed using four questions that addressed comfort, fear, and perception of effect on the consultation. Each question was assessed using a 5-point Likert scale, and all enrolled patients were asked to complete these questions.

#### CHW Experience Assessment

After the data collection period, a random subset of CHWs were invited to complete a survey and participate in focus group discussions and in-depth interviews to discuss perceived patient comfort, the effect of recording on consultation procedures, and the ease of managing recordings. Similarly, the survey questions focused on CHWs’ experience of being recorded, which was assessed using four questions that addressed ease, effects on focus, and perceived benefit. Each question was assessed using a 5-point Likert scale.

#### Translated Inputs

To compare model performance across languages, three-quarters (77.39%) of the encounter transcripts were randomly selected and translated into English using a combination of LLMs and professional linguists. These English transcripts were processed using the same prompts and procedures applied to the original Kinyarwanda transcripts. The outputs generated by the two large language models were then analyzed using the same analytical approach used for the original Kinyarwanda transcripts.

#### Sample Size

Initially, the study was intended to have two arms (1) CHWapp users, and (2) CHWs still using the paper version of the algorithms [M2]. However, the CHWapp was rolled out in the region selected for the paper-based arm during the data collection. Thus, this second arm was removed during the trial operationalization, and the study ended up focusing only on CHWapp users – the IRB was informed of the change.

The initial patient encounter sample size calculation was based on estimating the LLM’s sensitivity and specificity in making appropriate referral decisions, using methods adapted from Whittle et al. [M3] and informed by a benchmarking study that assessed multiple LLMs’ ability to answer medical questions posed by Rwandan CHWs [M4]. To detect a 20-percentage point difference in decision-making alignment with medical consensus (CHW vs. LLM), originally measured using a 5-point Likert scale [M4], and used as a proxy for referral decision accuracy (the primary outcome of this study), between 49 and 392 consultations (per arm, i.e., app-users or paper-users) was thought to be needed. These ranges assumed minimum cluster sizes of 7 interactions per CHW and accounted for clustering in the analysis.

Based on these projections, we planned to collect a total of 800 consultation transcripts, split evenly between two CHW cohorts: 400 recordings from CHWs using paper-based guidelines and 400 from CHWs using the CHWApp digital decision-support tool. To achieve this, we planned to recruit 50 CHWs per arm, each expected to contribute approximately 10 consultations. This recruitment target allowed for a 20% attrition rate due to technical failures, low recording quality, or CHWs submitting fewer than 7 recordings (the minimum cluster size for inclusion in analysis). We therefore anticipated that 40 CHWs per group would yield usable data, with an average of 10 transcripts per CHW, resulting in the required 400 consultations per arm. This sample size would have allowed for robust subgroup analyses and maintained adequate power to detect an absolute difference in referral accuracy as small as 11.8 percentage points—a conservative estimate given the larger effect sizes observed in benchmarking data

During the initial stages of data collection, it was noted that the attrition rate was much higher than expected (circa 70-80% observed at the start of the trial); as such, a decision was made to increase the number of CHWs recruited, with a revised target of 150, to achieve the patient encounter target. Moreover, a decision was made to drop the cluster minimum target and instead treat reaching 400 encounters as the stopping rule.

#### Statistical Analysis

A full statistical analysis plan was included with the original protocol [M2]. Descriptive statistics were used to summarize participant and consultation characteristics.

The primary analysis comprised two components:

1. Performance of CHWs & LLMs as determined against the expert panel-defined ground truth. Overall model performance, across all patients combined, was quantified using standard diagnostic measures, including accuracy, recall (sensitivity), specificity, precision (positive predictive value (PPV)), and negative predictive value (NPV), as mentioned above. Metrics, along with their corresponding 95% confidence intervals, were derived from a 2×2 confusion matrix for true positives (TP), false negatives (FN), false positives (FP), and true negatives (TN). In this context, a “positive” case represented a decision to refer, while a “true” case denoted that the decision to refer or not was deemed appropriate by the expert adjudication panel.
2. To compare the LLM’s performance with that of CHWs operating without LLM support, meta-analysis and meta-regression models were used. Initially, bivariate random-effects models were considered to jointly estimate correlated diagnostic measures while accounting for between-CHW heterogeneity. However, due to data sparsity, where approximately 70% of CHWs had no referral cases in their patient panels, random effects estimation was not feasible, as models either failed to converge or produced boundary estimates (**τ**² = 0). Consequently, separate one-stage meta-analysis models were fitted for each measure (accuracy, sensitivity, specificity, precision, and NPV), using fixed-effects binomial generalized linear models (GLMs) with a logit link function with evaluator (CHW, LLM1, LLM2) included as a three-level categorical predictor. This fixed-effects specification assumes no between-CHW heterogeneity in performance beyond sampling variability, which was necessary due to the convergence issues mentioned. Statistical differences in the CHW, LLM1 and LLM2 performances were determined from the fitted models, expressed as odds ratios (OR) and associated p-values derived from these comparisons, with uncertainty quantified using 95% confidence intervals (CI) calculated using Wald standard errors. A minimal continuity correction (0.01) was applied to confusion matrix cells to ensure numerical stability. Maximum likelihood estimation was used to fit all models. All analyses were performed in R utilizing the stats package.

All performance estimates were reported for both the LLM and CHW referral decisions, accompanied by 95% confidence intervals.

Other analyses include:

- The hindsight referral accuracy was analyzed using descriptive statistics and was reported as missed, unnecessary, or correct referral decisions.
- Likert scale responses assessing diagnosis and management quality were summarized descriptively, with frequency distributions presented in tables and central tendency summarized using the median. Histograms were used to visualize the proportion of cases rated at each scale level. Each outcome was also analyzed using a multilevel ordinal regression model that adjusts for key consultation characteristics, incorporating a random intercept to account for clustering by CHW.
- All of the analysis was replicated using translated English transcripts, and a meta-regression covariate was used to formally compare referral accuracy between the two languages.
- Patient and CHW user experience data were analyzed descriptively to evaluate the acceptability and perceived impact of consultation recordings.

Analyses were conducted in R version 4.5.2 [M5]. Data manipulation and visualization used the tidyverse package [M6]. Fixed-effects binomial generalized linear models were fitted using the stats package [M5]. Reporting followed the guideline for the Developmental and Exploratory Clinical Investigations of Decision Support Systems Driven by Artificial Intelligence (DECIDE-AI) [M7].

#### Qualitative Analysis

Thematic analysis was used to analyse the qualitative outputs from the CHW focus group and individual interviews. The translated transcripts were openly read independently by the two researchers. The researchers then met to create a codebook using Dedoose 10. Conflicting codes and codes that did not align with the study objectives were removed. The codes and excerpts identified through the inductive coding process were then collapsed into three main themes.

## Results

A comprehensive summary of the study’s activities and data flows is provided in Extended Data Figure 2.

### Study Cohort Characteristics

Of the 150 CHWs recruited, 126 CHWs had at least one eligible case in the final evaluation set, with 64 (50.8%) from Musanze and 62 (49.2%) from Nyabihu. Slightly more than half (n =74, 58.7%) of the CHWs were women, with similar distributions across districts. Ages ranged from 25 to 70 years (mean 44.8 ± 6.2), and most were between 45 and 54 years (n = 65, 43.7%). Education levels differed by district: 49 CHWs in Nyabihu (79.0%) had completed at least secondary education compared with 38 in Musanze (59.4%). CHWs had substantial experience, with a mean of 13.2 ± 6.2 years of service (Table 1).

**Table 1:** Baseline Characteristics of CHWs & Patients.

|  |  | Musanze n (%) | Nyabihu n (%) | Total N (%) |
| --- | --- | --- | --- | --- |
| **CHW Participants** |  |  |  |  |
| **Sample** |  | 64 (50.8%) | 62 (49.2%) | 126 |
| **Gender** | *Female* | 39 (60.94%) | 35 (56.45%) | 74 (58.73%) |
|  | *Male* | 25 (39.06%) | 27 (44.74%) | 52 (41.27%) |
| **Age (years)** | *25-34* | 5 (7.81%) | 8 (12.9%) | 13 (10.32%) |
|  | *35-44* | 29 (45.31%) | 17 (27.42%) | 46 (36.51%) |
|  | *45-54* | 24 (37.5%) | 31 (50.00%) | 55 (43.65%) |
|  | *≥55* | 6 (9.38%) | 6 (9.68%) | 12 (9.52%) |
| **Educational Status** | *Primary* | 26 (40.63%) | 13 (20.97%) | 64 (50.79%) |
|  | *Secondary and above* | 38 (59.38%) | 49 (79.03%) | 62 (49.21%) |
| **Patient Participants** |  |  |  |  |
| **Sample** |  | 218 (50.8%) | 211 (49.2%) | 429 |
| **Gender** | *Female* | 110 (50.5%) | 114 (52.3%) | 224 (52.21%) |
|  | *Male* | 108 (49.5%) | 97 (44.5%) | 205 (47.79%) |
| **Age (years)** | *0-5* | 198 (90.8%) | 197 (91.5%) | 391 (91.14%) |
|  | *≥6* | 20 (9.17%) | 18 (8.13%) | 38 (8.86%) |
| **Diagnosis** | *Gastroenteritis* | 108 (49.5%) | 87 (41.2%) | 195 (45.5%) |
|  | *Malaria* | 12 (5.5%) | 5 (2.4%) | 17 (4.0%) |
|  | *Pneumonia* | 25 (11.5%) | 39 (18.5%) | 64 (14.9%) |
|  | *URTI* | 64 (29.4%) | 70 (33.2%) | 134 (31.2%) |
|  | *Other* | 9 (4.2%) | 10 (4.7%) | 19 (4.4%) |
| **Recovery status** | *Recovered on day 3* | 197 (90.37%) | 164 (77.73%) | 361 (84.15%) |
|  | *Recovered on day 14* | 21 (9.63%) | 46 (21.80%) | 67 (15.62%) |

Between June 26 and September 28, 2025, the full CHW cohort recorded a total of 910 patient encounters, which equates to an average of 5 cases (IQR 3-8) per CHW. Of these, 404 were removed because they were incomplete (n = 256), inaudible (n = 56), or ineligible (i.e., consultations were not the first presentation of a patient’s clinical complaint) (n = 269). Some cases were excluded for multiple reasons. Another 54 cases were excluded due to missing follow-up data, 4 due to missing LLM outputs as Gemini 2.5 Flash refused to generate any outputs, flagging them as potentially dangerous or breaching protocol, and 19 due to data integrity issues caused by a technical error during case upload that resulted in some mislabeled cases. This left a final set of 429 cases for analysis (Figure 1), which were used to assess the decisions of CHWs and both LLMs.

**Figure 1:**
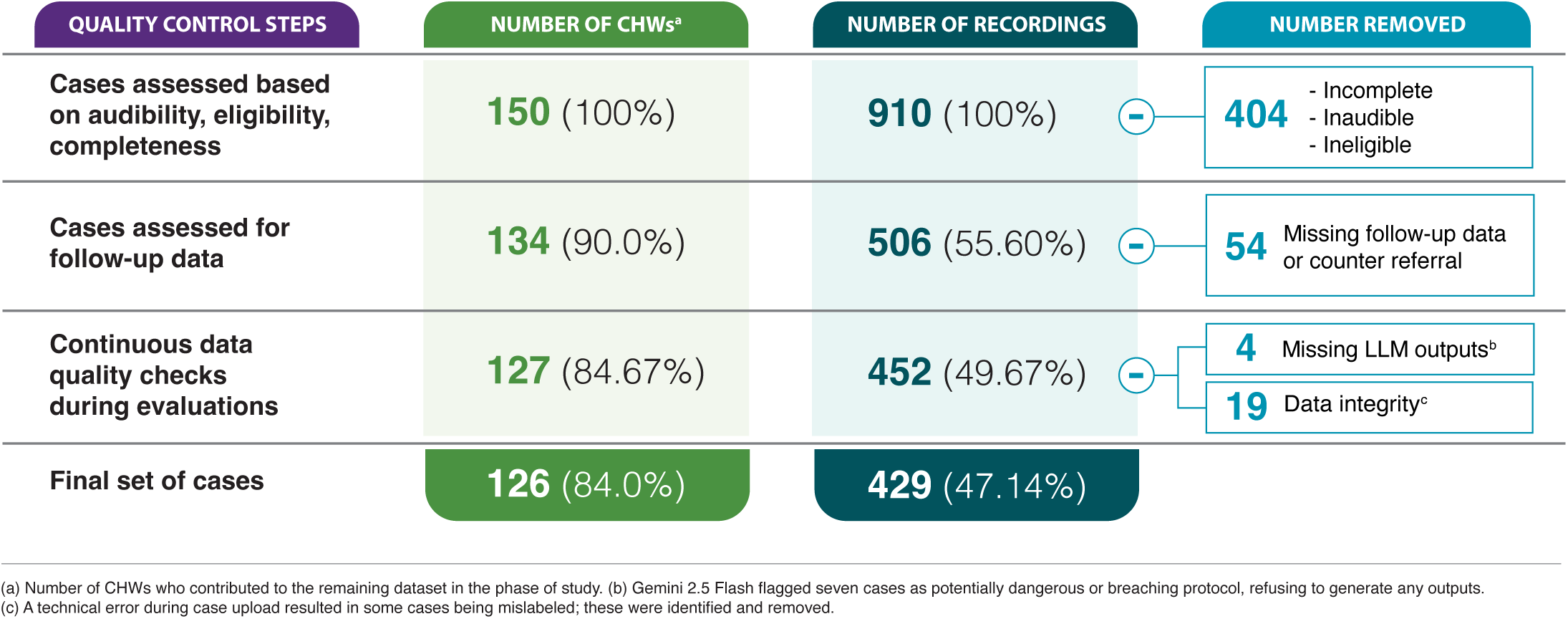
Quality control assessment of recordings and impact on sample size

Of the 429 cases analyzed, 218 (50.82%) were from Musanze and 211 (49.18%) from Nyabihu, and female patients represented a slight majority (224, 52.2%) across both sites. Nearly all patients were under 5 years old (n = 391, 91.1%), with only 38 (8.86%) aged 6 years or older. Most patients (n = 398, 92.77%) were not referred to health centers. Gastroenteritis and pneumonia accounted for more than three-quarters (n = 329, 76.69%) of the diagnoses. Most patients (84.15%) reported full recovery at day 3 follow-up, while one patient remained ill by the end of data collection (Table 1).

### Referral Accuracy

#### Initial-encounter referral accuracy

Based on the initial assessment data, the panel determined that 36 (8.39%) cases required referral. CHWs referred 31 cases; o3 referred 54 cases; and Gemini 2.5 Flash referred 262 cases. CHWs treated, without referral, 187 (95.9%) cases of gastroenteritis, 61 (95.31%) cases of pneumonia and 13 (76.47%) cases of malaria. Similarly, o3 declined to refer 177 (90.77%) cases of gastroenteritis, 55 (85.94%) cases of pneumonia and 14 (82.35%) cases of malaria. Gemini 2.5 Flash tended to refer many more cases, opting not to refer only 90 cases (46.15%) of gastroenteritis, 9 (14.06%) cases of pneumonia and 8 (47.06%) cases of malaria.

In comparison with ground-truth referral appropriateness (determined by the expert panel), CHWs demonstrated the highest accuracy (97.9% [95% CI: 96.1% - 98.9%)] and precision (93.5% [95% CI: 79.3% - 98.2%]), while o3 showed comparable accuracy (94.9% [95% CI: 92.4%-96.6%]), lower precision (63.0% [95% CI: 49.6%-74.6%]) and higher (absolute) recall (94.4% [95% CI: 92.4%-96.6%]), and Gemini 2.5 Flash had lower accuracy (47.3% [95% CI: 42.6% - 52.1%]) and precision (13.7% [95% CI:10.1%% - 18.4%]), due to its high false-positive rate (Table 2).

**Table 2:** Initial Referral Accuracy (Using Panel Decision as Reference)

|  | **CHW** | **Open AI o3** | **Gemini 2.5 Flash** |
| --- | --- | --- | --- |
| **True Positive** | 29 | 34 | 36 |
| **True Negative** | 391 | 373 | 167 |
| **False Positive** | 2 | 20 | 226 |
| **False Negative** | 7 | 2 | 0 |
| **Accuracy** | 97.9%<br>(96.1%-98.9%) | 94.9%<br>(92.4%-96.6%) | 47.3%<br>(42.6%-52.1%) |
| **Recall (Sensitivity)** | 80.6%<br>(65.0%-90.3%) | 94.4%<br>(81.9%-98.5%) | 100%<br>(90.4%-100%) |
| **Specificity** | 99.5%<br>(98.1%-99.9%) | 94.9%<br>(92.3%-96.7%) | 42.5%<br>(37.7%-47.7%) |
| **Precision (PPV)** | 93.5%<br>(79.3%-98.2%) | 63.0%<br>(49.6%-74.6%) | 13.7%<br>(10.1%-18.4%) |
| **NPV** | 98.24%<br>(96.4%-99.2%) | 99.5%<br>(98.1%-99.9%) | 100%<br>(97.8% - 100%) |

Table 3 presents comparative performance metrics across CHW and LLMs. In essence, CHWs demonstrated significantly higher accuracy compared to both LLMs. Gemini 2.5 Flash showed substantially lower odds of accurate classification relative to CHWs (OR = 0.02, p < 0.001; meaning Gemini-2.5-Flash was 50 times less likely than humans to produce accurate classification), while OpenAI o3 also performed significantly worse than CHWs, though with a smaller effect size (OR = 0.46, p = 0.031). In head-to-head comparison between LLMs, OpenAI o3 achieved significantly higher accuracy than Gemini 2.5 Flash (OR = 0.05, p < 0.001).

**Table 3:** Performance Comparison Between CHWs and LLMs using Fixed-Effects Binomial GLMs.

|  | Comparisons | OR [95% CI] | P value | Favors |
| --- | --- | --- | --- | --- |
| **Accuracy** | Gemini 2.5 Flash vs CHW | 0.02 [0.01-0.05] | < 0.001 | CHW* |
|  | OpenAI o3 vs CHW | 0.46 [0.22-0.93] | 0.031 | CHW* |
|  | Gemini 2.5 Flash vs OpenAI o3 | 0.05 [0.03-0.08] | < 0.001 | OpenAI o3* |
| **Recall (sensitivity)** | Gemini 2.5 Flash vs CHW | 8.07 [1.17-55.89] | 0.034 | Gemini 2.5 Flash* |
|  | OpenAI o3 vs CHW | 2.95 [0.75-11.63] | 0.122 | Not statistically significant |
|  | Gemini 2.5 Flash vs OpenAI o3 | 2.73 [0.33-22.48] | 0.349 | Not statistically significant |
| **Specificity** | Gemini 2.5 Flash vs CHW | 0.01 [0.00-0.02] | < 0.001 | CHW* |
|  | OpenAI o3 vs CHW | 0.15 [0.05-0.47] | 0.001 | CHW* |
|  | Gemini 2.5 Flash vs OpenAI o3 | 0.04 [0.03-0.07] | < 0.001 | OpenAI o3* |
| **Precision (PPV)** | Gemini 2.5 Flash vs CHW | 0.02 [0.01-0.06] | < 0.001 | CHW* |
|  | OpenAI o3 vs CHW | 0.18 [0.05-0.63] | 0.008 | CHW* |
|  | Gemini 2.5 Flash vs OpenAI o3 | 0.10 [0.05-0.19] | < 0.001 | OpenAI o3* |
| **NPV** | Gemini 2.5 Flash vs CHW | 2.81 [0.43-18.49] | 0.282 | Not statistically significant |
|  | OpenAI o3 vs CHW | 2.42 [0.67-8.78] | 0.180 | Not statistically significant |
|  | Gemini 2.5 Flash vs OpenAI o3 | 1.16 [0.15-9.16] | 0.886 | Not statistically significant |

Odds ratio (OR) interpretation: For "A vs B" comparisons, OR > 1 indicates A performs better; OR < 1 indicates B performs better
\*Statistically significant

Gemini 2.5 Flash demonstrated a higher sensitivity to refer with more than eightfold higher odds of correctly identifying patients requiring referral than CHWs (OR: 8.07, p = 0.034; though note this comes at the cost of a high false positive rate, see below). o3 achieved high sensitivity, and its performance did not differ significantly from that of CHWs (OR: 2.95, p = 0.122), nor was there a significant difference between the two LLMs (p = 0.349).

In contrast, CHWs consistently outperformed both LLMs in referral reliability, demonstrating significantly higher specificity and precision across all comparisons (p ≤ 0.008i). Gemini 2.5 Flash showed significantly lower odds to correctly identify patients not requiring referral relative to CHWs (OR: 0.01, p < 0.001), highlighting a pronounced sensitivity-precision trade-off. Gemini 2.5 Flash demonstrated significantly lower specificity (OR = 0.04, p < 0.001) and precision (OR = 0.10, p < 0.001) compared to OpenAI o3, indicating a greater tendency toward over-referral.

Despite these differences, no statistically significant variation was observed in negative predictive value (NPV) across all comparisons (all ps ≥ 0.180). Although Gemini 2.5 Flash achieved the highest raw NPV (99.3%), the already high baseline NPV among CHWs (97.9%) limited the potential for statistically detectable improvement.

#### Referral Accuracy (with more recent models)

During the course of the study, newer versions of the LLMs were released. Replication of the same referral appropriateness assessment for Gemini 3-pro and OpenAI GPT-5.1 yielded substantially improved results for Google’s LLM and slightly deteriorated OpenAI’s. Gemini 3-pro had an accuracy of 92.0% and a precision of 47.5%. GPT-5.1 had an accuracy of 94.1% and a precision of 56.3%. The two most notable shifts were that both models’ true positive rates deteriorated (Gemini 3-pro = 32, GPT-5.1 = 27), and whilst the OpenAI models’ false positive rate stayed almost the same (21 instances compared to 20 for the older model), the Google models’ false positive rate substantially decreased (32 instances compared to 226 for the older, smaller model).

#### Hindsight-Informed Referral Accuracy

After reviewing follow-up information, including counter-referrals when available, evaluators revised seven original referral decisions, changing them from referral to community management. When compared against the updated ground truth, LLM performance declined, whereas CHW performance improved (Table 4).

**Table 4:** Comparison of Initial Referral Decisions with Hindsight Referral Decisions.

| Referral decision | Gemini 2.5 Flash |  | OpenAI o3 |  | CHW |  |
| --- | --- | --- | --- | --- | --- | --- |
|  | Initial | Hindsight | Initial | Hindsight | Initial | Hindsight |
| Accurate referrals | 203 (47.3%,<br>95% CI:<br>42.6%-52.0%) | 198 (46.2%,<br>95% CI:<br>41.5% - 50.9%) | 407 (94.9%,<br>95% CI:<br>92.4%-96.6%) | 406 (94.6%,<br>95% CI:<br>92.1% - 96.4%) | 420 (97.90%,<br>95% CI:<br>96.1%-98.9%) | 425 (99.1%,<br>95% CI:<br>97.6% - 99.6%) |
| Unnecessary referrals | 226 (52.7%,<br>95% CI:<br>48.0%-57.4%) | 231 (53.9%,<br>95% CI:<br>49.1%-58.5%) | 20 (4.7%,<br>95% CI:<br>3.0%-7.1%) | 22 (5.1%,<br>95% CI:<br>3.4%-7.6%) | 2 (0.5%,<br>95% CI:<br>0.1%-1.7%) | 1 (0.2%,<br>95% CI:<br>0.0% - 1.3%) |
| Missed referrals | 0 | 0 | 2 (0.5%,<br>95% CI:<br>0.1%-1.7%) | 1 (0.2%,<br>95% CI:<br>0.0% - 1.3%) | 7 (1.6%,<br>95% CI:<br>0.8%-3.3%) | 3 (0.7%,<br>95% CI:<br>0.2% - 2.0%) |

### Differential Diagnosis Accuracy, Quality of Diagnostic Reasoning, & Management Plans

#### Diagnostic Accuracy (Extended Data Table 1)

OpenAI’s o3 had the panel’s reference diagnosis as its top-1 in 310 cases (72.3%), as its top-5 in another 100 cases (23.3%), and failed to list the ‘correct’ diagnosis in 19 cases (4.4%). On closer inspection, the most missed diagnosis by o3 was pneumonia, which was missed 8 times (12.5%) out of 64 cases of pneumonia in total (Figure 2a). Gemini 2.5 Flash had the panel’s reference diagnosis as its top-1 in 260 cases (60.6%), as its top-5 in another 135 cases (31.5%), and failed to list the ‘correct’ diagnosis in 33 cases (7.7%). On closer inspection, the most missed diagnoses for Gemini 2.5 Flash were gastroenteritis (9 cases, 4.61%) and pneumonia (11 cases, 17.1%; Figure 2a & Extended Data Table 2). Eight of the errors were common between the two LLMs (5 related to pneumonia, and one to each of: gastroenteritis, NCDs, and malaria).

**Figure 2A:**
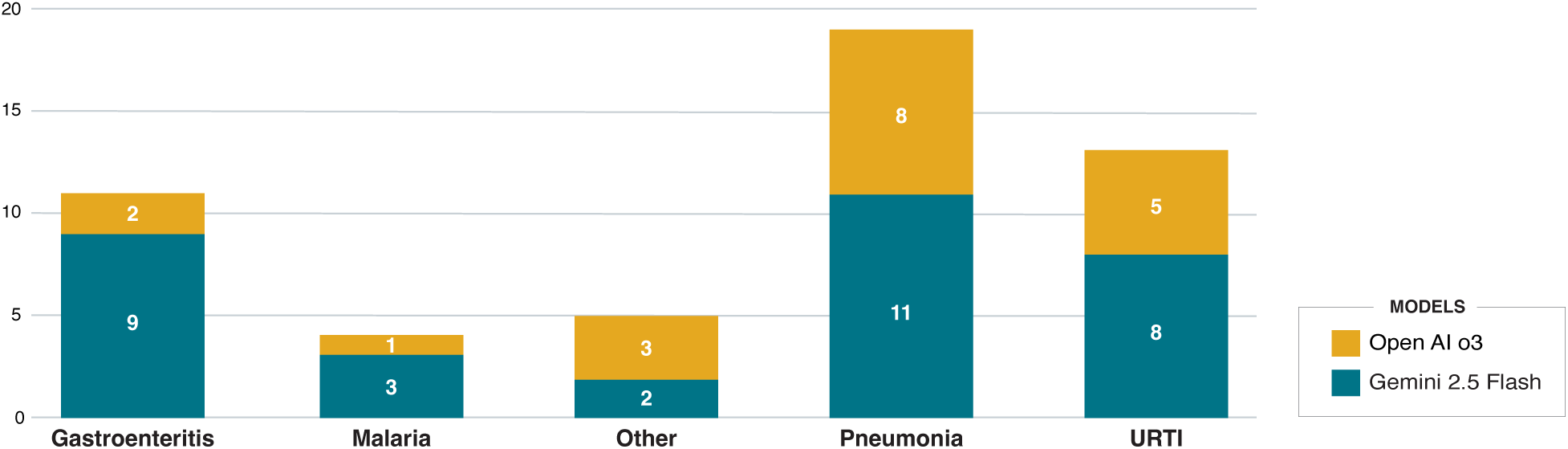
Frequency and type of diagnosis missed by the LLMs

**Figures 2B and 2C:**
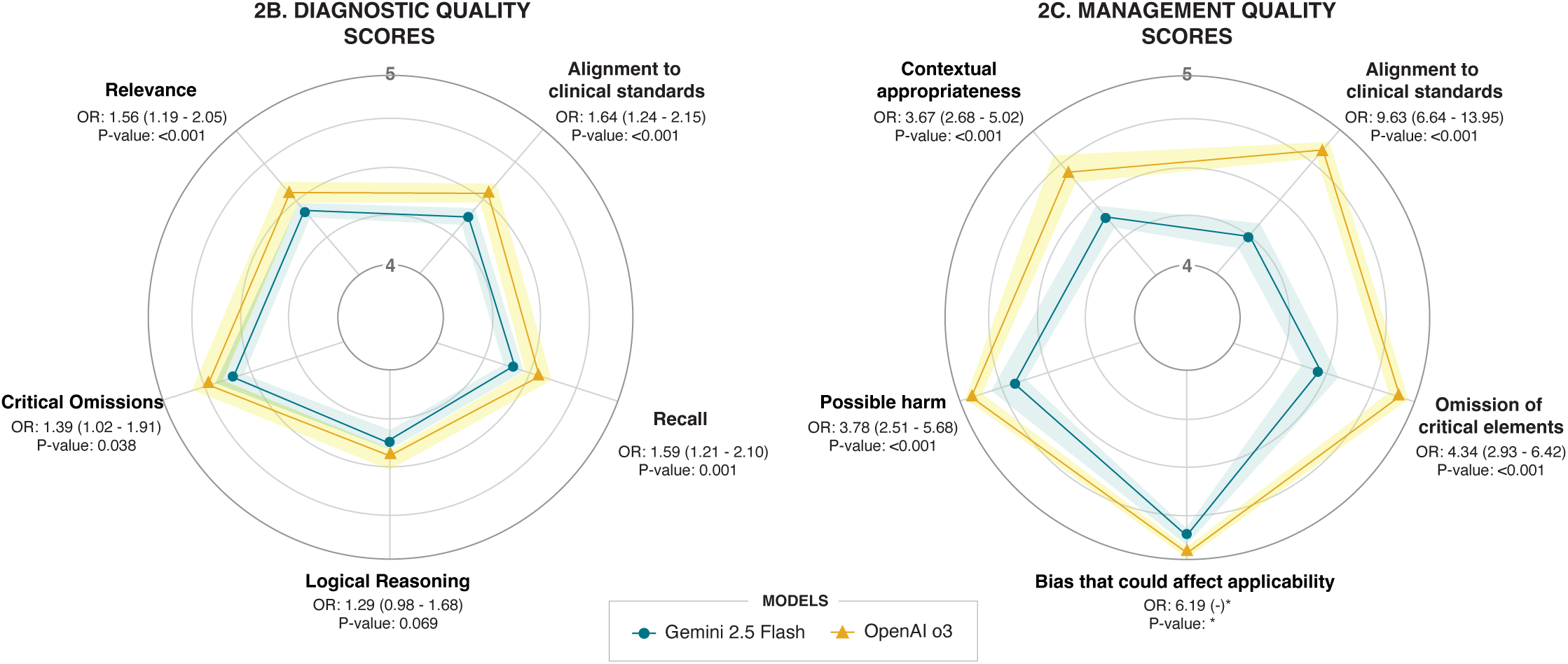
Quality scores for LLM outputs

#### Quality of Diagnostic Reasoning

OpenAI’s o3 demonstrated consistently superior performance relative to Gemini 2.5 Flash; o3 had a median score above 4.5, while Gemini 2.5 Flash maintained median scores above 4 across all diagnostic quality domains (Figure 2b). Notably, the differential diagnosis list from o3 was more likely to align with local standards (OR = 1.64; 95% CI: 1.24 – 2.15), more likely to include relevant differentials (referred to as ‘Recall’; OR = 1.59; 95% CI: 1.21 – 2.10), and less likely to miss critical conditions (OR =1.39; 95%CI: 1.02 – 1.91) or include irrelevant conditions (referred to as ‘Relevance’; OR = 1.56; 95%CI: 1.19 – 2.05), than Gemini’s list (Figure 2b).

#### Quality of Management Plan

Again, OpenAI’s o3 demonstrated consistently superior performance relative to Gemini 2.5 Flash; o3 had a median score above 4.5, while Gemini 2.5 Flash maintained median scores above four across all management plan quality domains. However, the effects were larger in the management plan domains than in the diagnostic reasoning domains. o3 showed markedly superior performance across all management safety domains, with substantially higher odds of alignment with clinical standards (OR = 9.63; 95%CI: 6.64 - 13.95), fewer important omissions (OR = 4.34; 95%CI: 2.93 - 6.42), and greater contextual appropriateness of management recommendations (OR = 3.67; 95%CI: 2.68 - 5.02). The potential bias metric was excluded due to a severe ceiling effect; 92% all ratings were 5/5, resulting in insufficient variation for reliable model estimation (Figure 2c).

When provided a binary decision of whether the management plans generated by the LLMs would have helped improve/guide more appropriate patient management had they been shown to the CHW, the evaluators judged that this was true in 78.32% (n = 335) of management plans generated by Gemini 2.5 Flash and 91.14% (n = 391) of those generated by o3. Finally, assessment of the appropriateness of the language utilized by the LLMs revealed that both models consistently employed medical language suitable for CHW comprehension. OpenAI o3 achieved 100% appropriate terminology usage while Gemini 2.5 Flash demonstrated 99.3% (n = 426) appropriateness.

#### Language-related Insights

Analysis of translated (to English) outputs confirmed o3’s superior performance. In English, o3 significantly outperformed Gemini 2.5 Flash in specificity (97.7% vs 47.7%, p<0.0001) and precision (75.9% vs 14.6%, p=0.004). No statistically significant differences in performance were observed between the English and Kinyarwanda outputs for either model (Extended Data Table 3). Negligible heterogeneity (**τ**²<0.001) indicated consistent performance across CHW clusters.

### User-Centered Quantitative & Qualitative Insights

#### CHW (User)-centered Insights

40 CHWs were included in the post-data collection user experience study. When asked how often patients expressed concerns about being recorded: 20 (50.0%) stated ‘Never’, 14 (35.0%) ‘Rarely’, and 6 (15.0%) stated ‘Sometimes’. More than half (n = 24, 60.0%) reported that recording did not at all influence their consultation, 4 (10.0%) reported mild influence, while 7 (17.5%) reported moderate influence. Asked about how much recording affected their ability to focus on patient care, more than half (n = 22, 56.41%) reported ‘Not at all’, while 10 (25.64%) reported ‘Moderate’ effect on focus. One CHW mentioned that recording extremely affected their focus during consultation. Additionally, all CHWs indicated that additional training would be beneficial.

To further understand CHW’s experience, in-depth interviews and focus group discussions were conducted. The outcomes mapped to three major themes: an initial apprehension about being recorded, a subsequent change in behaviour and communication practices, and CHWs’ perceptions of the patient experience.

Initially, many CHWs experienced considerable discomfort with the recording process. The knowledge that their voices would be captured and potentially reviewed created feelings of vulnerability and exposure. Some CHWs expressed fear about forgetting important information or saying something inappropriate during consultations. This anxiety was particularly pronounced in the early days of implementation.

> “At first the recording scared me…I thought maybe I would say something wrong.” (M12, Musanze)

However, these negative emotions gradually waned as CHWs became more familiar with recording. The repetition of recording sessions, combined with supportive training, helped shift their perspective. This acceptance is evident in statements from CHWs who noted they no longer even thought about the phone recording them anymore.

> “Now I am used to it…I don’t even think about the phone anymore.” (N29, Nyabihu) “Recording made me more confident…” (N52, Nyabihu)

The second theme concerned changes in how CHWs conducted consultations. They expressed that being recorded influenced them to become more deliberate and careful in their communication. Many reported paying closer attention to their words, speaking more clearly, organizing their thoughts more carefully, and following proper procedures. This heightened attention to quality suggests that recording catalyzed more professional and structured interactions with patients.

> “Recording helped me be more careful and speak clearly.” (N52, Nyabihu)

> “Recording reminded me to focus and not forget important things.” (M12, Musanze)

CHWs described a range of patient responses from initial concern to acceptance once the purpose was explained. A small number of patients were hesitant at first, but almost all patients agreed when they understood it was to improve service quality.

> “Some asked why we are recording…they wanted to know the reason first.” (M07, Musanze)

> “When I told them it was to help us do better, they agreed.” (N11, Nyabihu)

Patient hesitation was mostly due to unfamiliarity. Their questions show a desire to understand the purpose rather than resistance. Acceptance grew quickly when patients understood the intention behind recording.

#### Patient-Centered Acceptability Insights

All patients completed the user experience survey. Most reported feeling able to speak freely during their consultations, with 107 (24.95%) agreeing (score of 4 out of 5) and 321 (74.83%) strongly agreeing (score of 5 out of 5). Similarly, most felt comfortable with their consultation being recorded: 86 (20.05%) agreed (score of 4 out of 5), 338 (78.79%) strongly agreed (score of 5 out of 5), and only 5 (1.17%) reported discomfort (score of either 1 or 2). Patients also commonly perceived the consultation as more thorough due to the recording, with 105 (24.48%) agreeing (score of 4 out of 5), and 324 (75.52%) strongly agreeing (score of 5 out of 5). Nearly all indicated that the recording process did not negatively affect the evaluation process, with 114 (26.58%) agreeing (score of 4 out of 5), and 315 (73.42%) strongly agreeing (score of 5 out of 5).

## Discussion

This study provides the first real-world assessment of the potential accuracy of ‘ambient listening’ in combination with LLMs for improving the quality of community-based healthcare in a low-resource setting. By analyzing 429 clinical encounters captured during routine service delivery, we show substantial differences in how two state-of-the-art LLMs perform on critical tasks such as referral decision-making, differential diagnosis, and management plan development. The very high baseline performance of the CHWs makes the performance of the LLMs less immediately encouraging; however, in context, this current generation of models demonstrates clear promise, and their behaviour illustrates key considerations for safe, context-appropriate deployment. For example, in contrast to o3’s overall strong performance, Gemini 2.5 Flash’s pattern of referring nearly every patient, illustrates the core challenge of using models that default toward risk minimization under uncertainty – a behavior well-documented in LLMs trained with reinforcement learning from human feedback (RLHF) frameworks that heavily penalize harmful omissions, and manifest as exaggerated safety responses, leading models to refuse valid requests (as noted with Gemini but not o3), or exhibit an overcautious disposition [25,26]. This overcautiousness, while intended to prevent harmful outputs, inadvertently reduces the practical usefulness and trustworthiness of LLMs in nuanced clinical settings, as they encounter frequent, unnecessary referrals or rejections of valid diagnostic pathways [27]. At scale, excessive referrals can strain primary care facilities and reduce trust among CHWs [28,29], and thus, in terms of clinical usefulness, the results of this study point to a clear ‘winner’ in the head-to-head comparison of the two LLMs.

### In Context of the Literature

The markedly lower referral rate observed in this study (7.23%), compared to prior knowledge (circa 48.7%) which was incorporated into the study design [30], can be explained through a number of different means. For example, the extensive experience of the CHWs recruited (average ∼a decade) and the subsequent repeated exposure to the small set of well-defined conditions that made up the majority of cases recorded could have reduced CHW cognitive load and reinforced their understanding of how to manage many of the cases encountered (in line with established algorithms) [31,32]. But more importantly, the result exemplifies the previously reported finding that CHWs, when supported by structured algorithms, tend to make balanced and contextually appropriate decisions that minimize unnecessary referrals [33,34]. This is critically important for identifying situations in which LLMs might have the greatest impact, i.e., LLM-based CDSS might be more effective in situations that lack formal, structured algorithms at baseline and have less experienced CHW cohorts (due to cohort expansion, turnover, the addition of novel responsibilities, etc.).

The strong diagnostic reasoning capabilities demonstrated by both models, as evidenced by their frequent identification of the correct diagnosis among their top-one and top-five differential diagnoses, are consistent with several prior studies [21,35,36]. However, the models exhibit notable limitations, missing over 10% of pneumonia diagnoses. Pneumonia remains one of the leading causes of pediatric mortality in many low-resource settings. Such diagnostic errors in identifying pneumonia can delay early detection and lead to poor clinical outcomes [37,38]. Moreover, the demonstration that translation to English before inputting text into the LLMs yields better diagnostic and management plan-related outcomes, is concordant with prior studies both in Kinyarwanda [21], and other African languages [39], reinforcing the need for further investment in language capabilities.

The user experience data offered significant insights into the acceptability of ambient listening during CHW clinical encounters. CHWs’ initial discomfort, manifested as feelings of vulnerability and apprehension about errors, aligns closely with the broader literature, which attributes healthcare providers’ concerns about professional identity, fear of exposure of skills, and a perceived loss of control when introduced to recordings in their practice [40]. Nonetheless, their subsequent adaptation and growing confidence exemplify a prevalent pattern wherein providers’ initial reluctance subsides through familiarity and appropriate support, often yielding improved performance [40–42]. The combined evidence of patients’ overwhelmingly positive feedback, evidenced by high comfort and confidence in recorded sessions, and CHWs’ successful acclimation implies that, despite early challenges, recording clinical encounters can deliver net benefits with adequate training and support. This alignment provides compelling evidence for the feasibility and acceptability of embedding ambient recording in CHW workflows to allow the potential integration of LLM-based CDSS tools.

### Strengths & Limitations

The key strength of the study is that it was carried out based on real-world clinical encounters rather than simulated cases, providing ecological validity that simulated studies cannot achieve. Moreover, our analysis accounted for the hierarchical structure of the data, reducing the risk of biased or misleadingly precise performance estimates. Finally, the use of multiple evaluators and structured quality assessments reduced individual bias and improved the reliability of performance judgments. Despite the study’s strengths, the results must be viewed in light of certain limitations. Even though consultations were recorded silently and without the investigator’s presence, the fact that patients were informed and consented to be recorded means that there remains a non-trivial risk of a Hawthorne effect. Additionally, although CHWs receive training across multiple domains, including family planning, nutrition, and non-communicable diseases, their day-to-day patient-initiated clinical work focuses mainly on malaria, pneumonia, malnutrition, and gastroenteritis in children under five [43,44]. While this reflects the current operational scope of the integrated community case management (iCCM) program in Rwanda [45,46], it limits the generalizability of the results and may lack the power to identify more subtle variations in safety or effectiveness across clinical subgroups. Furthermore, the low referral rates further constrained the precision of our model comparisons and reduced the statistical power to detect meaningful differences. Lastly, because certain safety nets were implemented to prevent data loss (e.g., capture and storage locally on the device rather than real-time transmission using cellular signal), we cannot provide operational insight to inform future deployments.

### Implications for Policy Makers, Product Developers & Researchers

Model selection for this study was informed by a Rwandan CHW-specific benchmarking exercise that evaluated multiple LLMs on clinical reasoning, knowledge recall, and alignment with local context [21] and identified predecessors (i.e., older, smaller versions) of the models used in this study as being able to provide superior advice to local primary care physicians. The critical insight that emerges from comparing the benchmarking results with those from this study is that in-silico evaluations do not appear to provide strong guarantees of future in-field performance on related (but not identical) tasks. Alternatively, the results could be attributed to the Likert-based assessment of ‘content quality’ not being predictive of performance on specific clinical tasks such as referral accuracy. The key implication is a stark warning against over-reliance on in silico evidence as justification for a model’s readiness for field deployment, especially when tasks are non-identical.

That said, assuming the right LLM is selected, this study’s findings suggest that this workflow can yield referral decisions with an accuracy comparable to that of highly experienced CHWs (average > 10 years of experience), which could be useful in the context of supportive supervision for less experienced CHWs. Studies have shown that expanding CHW work packages without adequate support can lead to decreased performance and effectiveness [10,47], which a version of the proposed workflow could prevent. However, a critical concern that this potential needs to be balanced against is automation bias – the tendency for human users to over-rely on automated recommendations, even when those recommendations are incorrect [48–50]. CHWs working with limited supervision, heavy caseloads, and little medical training may be particularly vulnerable to this cognitive bias [51]. If CHWs blindly trusted LLM outputs without critical evaluation, both under-referral (from a model like OpenAI o3, which occasionally misses serious conditions) and over-referral (from a model like Gemini 2.5 Flash) could compromise patient safety and the health system’s functioning. Whilst beyond the scope of this study, the impact of automation remains a key issue for future research, as it is likely that user interface design choices can be used to facilitate a learning effect rather than a deskilling one [52].

### Future Research

Given that earlier-stage evidence provides few guarantees (especially in this study, where many operational challenges have been circumvented by the design), a future research study might examine how CHWs interact with a similar workflow via an interventional trial, ideally integrating the LLMs directly into the CEMR to assess real-time usability, safety, and impact on both health system efficiency and patient-level outcomes. Additionally, this study highlights several other questions that warrant further exploration. The LLMs in this study were operationalized with robust prompts, but no anchoring knowledge (i.e., without formal access to Rwandan CHW guidelines). It is entirely possible that the performance of both LLMs could be improved using a RAG framework. In the case of o3, even a marginal improvement could make the tool statistically significantly better than the CHWs. Furthermore, recent advances have shown the ability of ambient scribes to reduce cognitive burden and burnout risk amongst clinicians [53]; it would be worth exploring the added value of LLM-based summarization of the transcripts collected, as summary documentation is not something that CHWs in Rwanda currently produce. By incorporating these summaries into the referral workflow, the quality of care delivered downstream could be improved through more appropriate patient triage and management [54,55], reflecting an alternative strategy by which the ‘ambient listening’ plus LLM workflow could add operational and clinical value.

## Conclusion

In summary, this ‘silent trial’ did not demonstrate the superiority of LLMs over CHWs in making correct referrals, but it provides emerging evidence that some LLMs have the potential to be safe and effective as clinical decision support systems for frontline health workers. In particular, the trial shows that the choice of LLM is hugely important, with o3 but not Gemini 2.5 Flash demonstrating referral accuracy and clinical reasoning comparable to those of highly experienced CHWs; however, more recent and larger versions of the Gemini model (i.e., ‘version 3 pro’) were much closer to equivalence. Additionally, more investment is still needed in the enabling environment before large-scale roll-out can be justified; from robust training to prevent automation bias [48], clear protocols for human oversight, continuous performance monitoring, and governance frameworks that ensure accountability and safety – much of which is missing in the contexts that have the most to gain from these tools.

## Data Availability

The collected data includes identifiable information such as initials, addresses, and affiliated health centers. Due to the sensitive nature of this information and in compliance with ethical standards, it is not feasible to provide a public dataset. However, the data can be made available by the University of Global Health Equity (UGHE) Institutional Review Board (IRB), under conditions that ensure the confidentiality and protection of participants' rights. The UGHE IRB can be contacted via email at irb.ughe.org or Kigali Heights, Plot 772. KG 7 Ave., 5th Floor, PO Box 6955, Kigali, Rwanda. Access to the data will be controlled in accordance with the requirements of the Rwandan government. Applications for data will be reviewed by a joint committee comprising representatives from the Rwanda Biomedical Centre and the University of Global Health Equity.

## Author contributions (CRediT taxonomy)

Conceptualization: BAM; Methodology: VM, BAM, MEF, NS, XL; Software: SR; Data curation (extraction, de-identification, management): AU, NS, MK; Investigation: all authors; Formal analysis: MK; Visualization: MK; Project administration: MEF, CN; Supervision: NS, BAM; Funding acquisition: BAM; Writing (original draft): NS, RW, SR, IR, BAM; Writing (review and editing): all authors

## Acknowledgments

We express our deep appreciation to the Community Health Workers whose recordings, consistent follow-up, and commitment were essential to this study. We also gratefully acknowledge the contributions of Samuel Habimana, Alliance Umuhoza, Saad Byiringiro, Mivumbi Michael Patrick, Francis Nkurunziza, Claude Kwizera, and Gwydion Williams, each of whom provided valuable support at different stages of the research process.

## Ethics & Data Management

The study protocol was reviewed and approved by the Rwanda National Ethics Committee (RNEC 853/2025). The study was prospectively registered in the Pan-African Clinical Trials Registry (PACTR202504601308784). All electronic data, including audio files, transcripts, annotations, LLM outputs, and evaluator ratings, was encrypted and stored on secure servers with controlled access. Physical documents, including consents and site files, were archived in locked facilities in accordance with institutional policy. Data quality was maintained through automated validation checks within the data system, manual review of transcripts, and periodic audits.

---

**Extended Data Figure 1:**
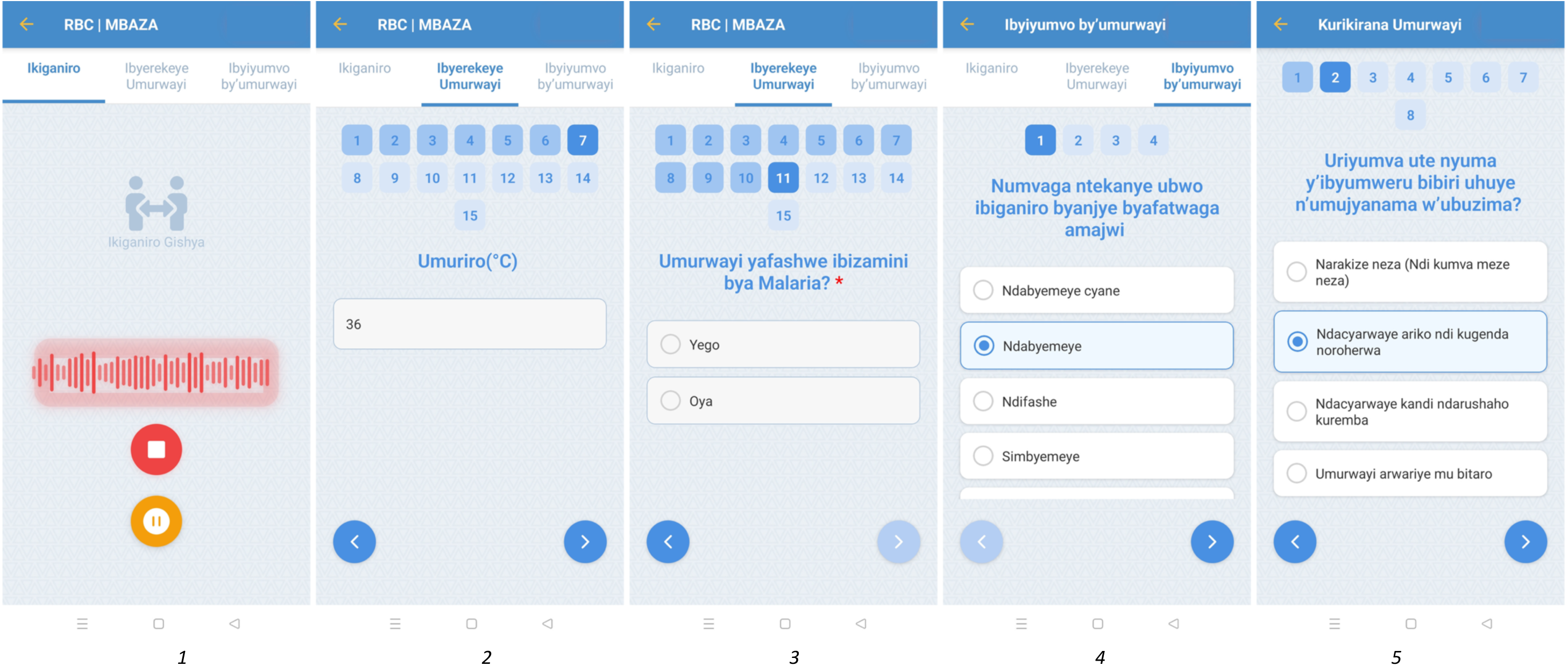
The Mbaza Application (1) First image on the left displays the ambient recording of the conversation between the CHW and the patient. (2 and 3) show the user interface of the app used to take patient information including demographics, vital signs (when applicable), Malaria RDT results and whether the patient was referred to the health center, in total there are 15 questions asked (3) The fourth image show the user interface for the patient experience questionnaire (5) The last image depicts the follow up questions interface used to capture information during the day 3 and day 14 follow up visits.

**Extended Data Figure 2:**
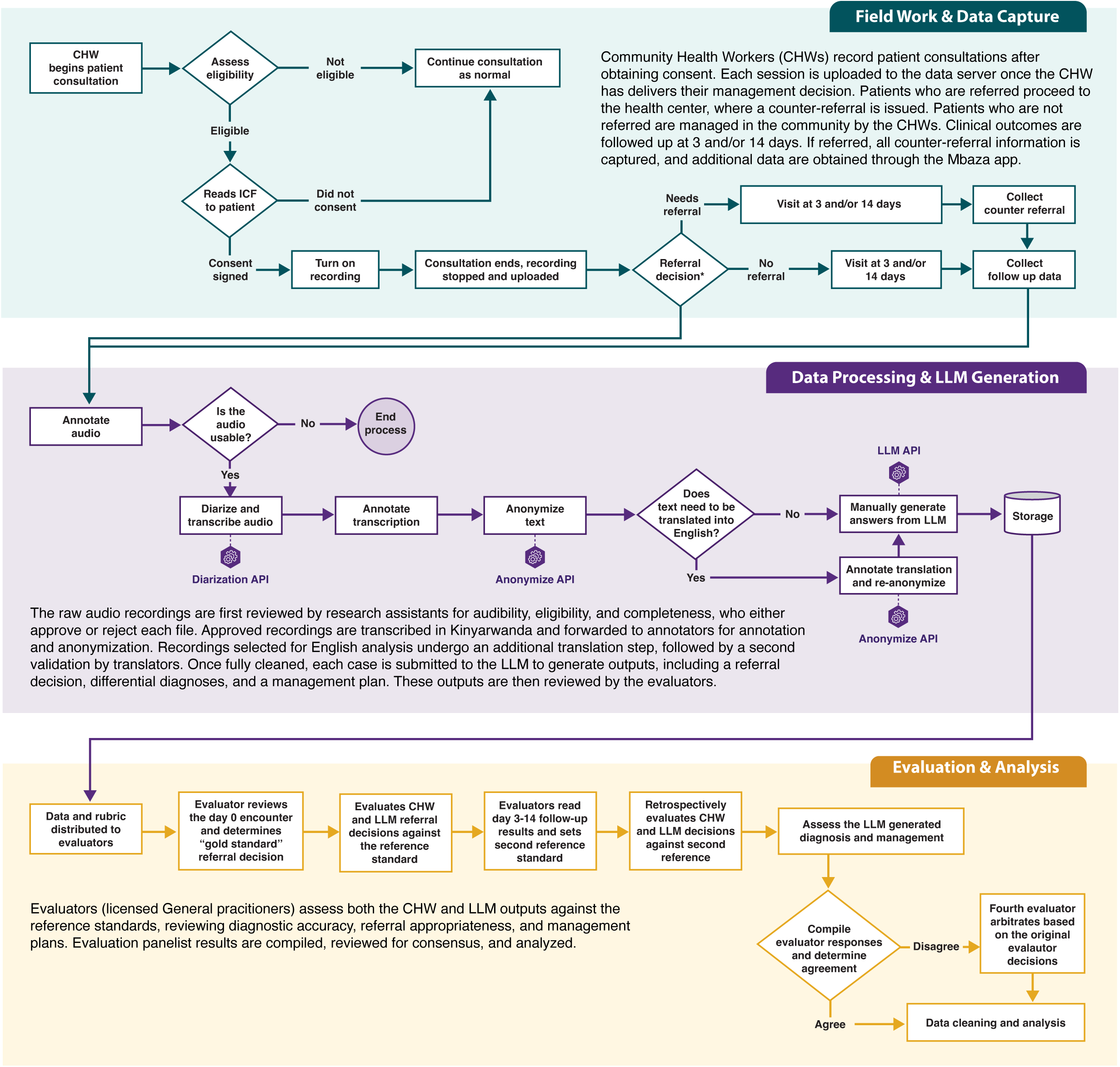
Trial Activities and Data Flow

**Extended Data Table 1:**
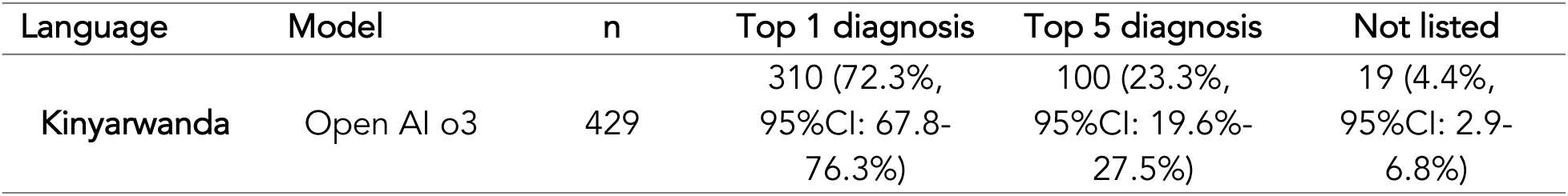
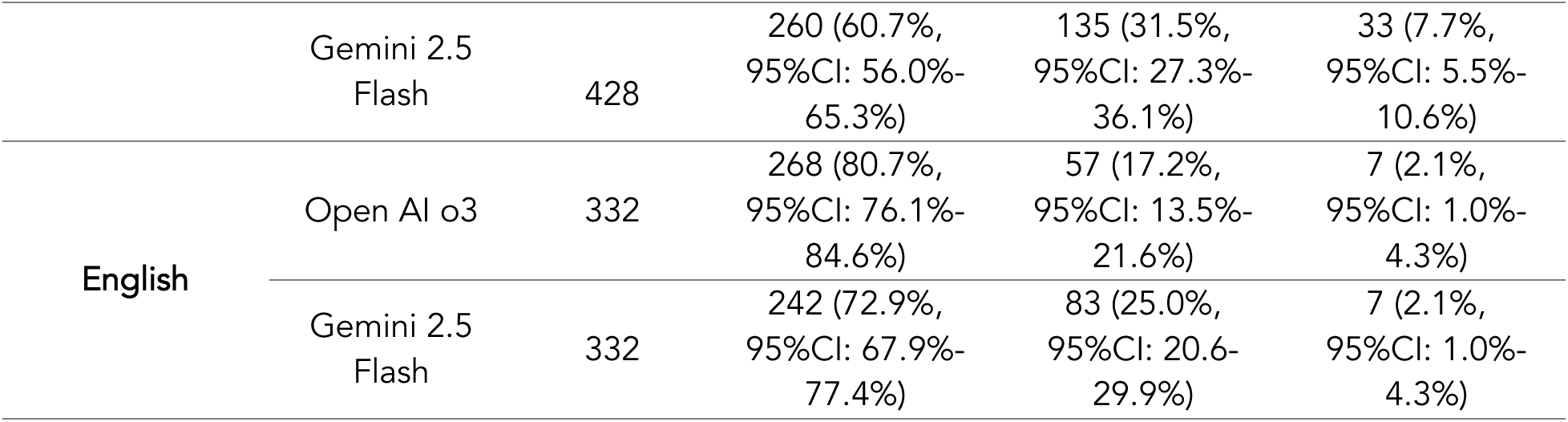
Diagnostic Accuracy Descriptive Statistics.

**Extended Data Table 2:** Non-Referrals Per Diagnosis Per Language.

| Diagnosis | Total Cases | Gemini 2.5 Flash (Kinyarwanda) | Gemini 2.5 Flash (English) | OpenAI o3 (Kinyarwanda) | OpenAI o3 (English) | CHW |
| --- | --- | --- | --- | --- | --- | --- |
| Gastroenteritis | 195 | 90<br>46.2% (39.3%-53.2%) | 70<br>35.9% (29.5%-42.8%) | 177<br>90.8%<br>(85.9%-94.1%) | 133<br>68.2%<br>(61.4%-74.3%) | 187<br>95.9%<br>(92.1%-97.9%) |
| URTI | 134 | 57<br>42.5% (34.5%-51.0%) | 55<br>41.0%<br>(33.1%-49.5%) | 125<br>93.3%<br>(87.7%-96.4%) | 110<br>82.1%<br>(74.7%-87.7%) | 132<br>98.5%<br>(94.7%-99.6%) |
| Pneumonia | 64 | 9<br>14.1% (7.6%-24.6%) | 13<br>20.3% (12.3%-31.7%) | 55<br>85.9%<br>(75.4%-92.4%) | 44<br>68.8%<br>(56.6%-78.8%) | 61<br>95.3%<br>(87.1%-98.4%) |
| Malaria | 17 | 8<br>47.1% (26.2%-69.0%) | 7<br>41.2% (21.6%-64.0%) | 14<br>82.4%<br>(59.0%-93.8%) | 9<br>52.9%<br>(31.0%-73.8%) | 13<br>76.5%<br>(52.7%-90.4%) |
| Others* | 19 | 3<br>15.8% (5.5%-37.6%) | 2<br>10.5% (2.9%-31.4%) | 4<br>21.1% (8.5%-43.3%) | 7<br>36.8%<br>(19.1%-59.0%) | 5<br>26.3%<br>(11.8%-48.8%) |

\*Other included diagnosis such as dermatological complaints, headache, and myalgias.

**Extended Data Table 3:** Language Comparisons for Gemini 2.5 Flash And o3.

|  | **Gemini 2.5**<br>**Flash (Kinyarwanda)** | **Gemini 2.5**<br>**Flash (English)** | **Difference** | **p- value** |
| --- | --- | --- | --- | --- |
| **Accuracy** | 47.9%<br>(42.6%-53.3%) | 51.5%<br>(46.1%-56.8%) | -0.04<br>(-0.08-0.02) | 0.20 |
| **Recall**<br>**(sensitivity)** | 100.0%<br>(88.6%-100.0%) | 90.0%<br>(74.4%-96.5%) | 0.10<br>(0.00-0.22) | 0.24 |
| **Specificity** | 42.7%<br>(37.3%-48.4%) | 47.7%<br>(42.1%-53.3%) | -0.05<br>(-0.10-0.01) | 0.10 |
| **Precision (PPV)** | 14.8%<br>(10.6%-20.3%) | 14.6%<br>(10.2%-20.4%) | 0.00<br>(-0.02-0.02) | 0.89 |
| **NPV** | 100.0%<br>(97.1%-100.0%) | 98.0%<br>(94.2%-99.3%) | 0.02<br>(0.00-0.05) | 0.23 |
|  | **Open AI o3 (Kinyarwanda)** | **Open AI o3 (English)** | **Difference** | **p- value** |
| **Accuracy** | 94.6%<br>(91.6%-96.5%) | 95.5%<br>(92.7%-97.2%) | -0.01<br>(-0.04-0.02) | 0.64 |
| **Recall**<br>**(sensitivity)** | 93.3%<br>(78.7%-98.2%) | 73.3%<br>(55.6%-85.8%) | 0.20<br>(0.06-0.35) | 0.02 |
| **Specificity** | 94.7%<br>(91.6%-96.7%) | 97.7%<br>(95.3%-98.9%) | -0.03<br>(-0.05- -0.01) | 0.02 |
| **Precision (PPV)** | 63.6%<br>(48.9%-76.2%) | 75.9%<br>(57.9%-87.8%) | -0.12<br>(-0.26-0.01) | 0.09 |
| **NPV** | 99.3%<br>(97.5%-99.8%) | 97.4%<br>(94.9%-98.7%) | 0.02<br>(0.01-0.04) | 0.02 |
| difference > 0 : in favor of Google's Gemini 2.5-Flash/Open AI o3 (Kinyarwanda) |  |  |  |  |
| difference < 0 : in favor of Google's Gemini 2.5-Flash/Open AI o3 (English) |  |  |  |  |

